# Metabolic subgroups and cardiometabolic multimorbidity in the UK Biobank

**DOI:** 10.1101/2021.02.01.21250893

**Authors:** Anwar Mulugeta, Elina Hyppönen, Mika Ala-Korpela, Ville-Petteri Mäkinen

## Abstract

**Background:** 

Ischemic heart disease (IHD), diabetes, cancer and dementia share features of age-associated metabolic dysfunction. We hypothesized that metabolic diversity explains the diversity of morbidity later in life.

**Methods:** We analyzed data from the UK Biobank (N = 329,908). A self-organizing map (SOM, an artificial neural network) was trained with 51 metabolic traits adjusted for age and sex. The SOM analyses produced six subgroups that summarized the multi-variable metabolic diversity. The subgroup with the lowest adiposity and disease burden was chosen as the reference. Hazard ratios (HR) were modeled by Cox regression (P < 0.0001 unless otherwise indicated). Enrichment of multi-morbidity over random expectation was tested by permutation analysis.

**Results:** The subgroup with the highest sex hormones was not associated with IHD (HR = 1.04, P = 0.14). The subgroup with high urinary excretion without kidney stress (HR = 1.24) and the subgroup with the highest apolipoprotein B and blood pressure (HR = 1.52) were associated with IHD. The subgroup with high adiposity, inflammation and kidney stress was associated with IHD (HR = 2.11), cancer (HR= 1.29), dementia (HR = 1.70) and mortality (HR = 2.12). The subgroup with high triglycerides and liver enzymes was at risk of diabetes (HR = 15.6). Paradoxical enrichment of multimorbidity in young individuals and in favorable subgroups was observed.

**Conclusions:** These results support metabolic diversity as an explanation to diverging morbidity and demonstrate the potential value of population-based metabolic subgroups as public health targets for reducing aggregate burden of chronic diseases in ageing populations.

**Key messages:**

- We introduced six data-driven subgroups of the UK Biobank as a high-dimensional model of metabolic diversity and disease risk within a human population.
- Three subgroups captured features of the classical cardiometabolic spectrum with stratification along cholesterol and blood pressure, kidney or liver dysfunction and systemic inflammation.
- Two novel subgroups of high sex hormones and high urinary excretion were observed.
- We defined a new concept of multimorbidity enrichment.
- nexpected patterns of multimorbidity indicated that metabolically “healthy” individuals with one cardiometabolic disease may be at a disproportional synergistic risk of co-morbidity.

## Introduction

The top 10 global causes for death included ischemic heart disease (IHD, 1st), stroke (2nd), dementias (5th), respiratory cancers (6th) and diabetes (7th) according to the Global Health Estimates 2016 report by the WHO. Much of this disease burden is attributed to obesity-associated metabolic dysfunction that increases the risk of cardiometabolic diseases^1^, multiple cancers^2^ and dementia^3^ in ageing individuals. These associations are supported by experimental studies of ageing^4^. There is thus a causal rationale why population subgroups with poor metabolic health bear a higher aggregate burden of multiple chronic diseases later in life.

The predictive power of metabolic profiling has been demonstrated in human populations^5,6^, yet the practical value may be limited for an individual patient^7–9^. In fact, the juxtaposition between individual and population health is a fundamental tenet of epidemiology and the two domains may never be reconciled simultaneously^10,11^. We propose data-driven subgrouping as a complementary solution that combines the robustness of population-wide statistics while retaining direct connections to observable individual metabolic profiles^12,13^.

In a typical scenario, people with similar profiles are grouped together and the aggregate rates of disease outcomes are compared between the subgroups. Examples of biomarker-based analyses include five clusters of diabetes with divergent outcomes^14^, four metabolic profiles of cancer mortality^15^ and six endotypes of heart failure^16^. We developed subgroups of diabetic complication burden in 2008^17^ and validated them in 2018 with new previously unseen data on clinical outcomes^18^. These studies are highly valuable since they produce quantitative descriptors of population health (subgroup profiles) that contain clues on how to reduce adverse long-term outcomes (biological interpretation of the biomarker profiles).

The UK Biobank includes half a million participants, 51 anthropometric and biochemical variables and ten years of follow-up data. Thus, it provides a unique opportunity to investigate relationships between the quantitative descriptors of metabolism and the development of morbidities^19^. To bridge the gap between health of a population as a whole and the individuals therein, we introduce the self-organizing map (SOM^13^) of the UK Biobank. Our results show how the health of an entire population can be summarized using an artificial neural network and how the subgrouping concept yields new insights into public health as more and more high-dimensional medical data become available.

## Materials and Methods

The UK Biobank is a prospective cohort study of over 500,000 participants aged 37-73 years recruited between 2006 and 2010^19^. Participants provided baseline information, physical measures and blood and urine samples and information on disease outcomes was obtained through register linkage, including Hospital Episode Statistics (HES), cancer and national death registries. The dataset included in this study comprised 153,731 men and 176,177 women of white British ancestry (Supplementary Figure S1).

The self-organizing map (SOM) is an artificial neural network approach that is designed to facilitate the detection of multi-variable patterns in complex datasets^20^. The result of the analysis is a two-dimensional layout where individuals with similar profiles are close together on the map and thus can be assigned to the same subgroup by visually observable proximity. In this respect, the SOM is a type of clustering analysis, however, in our framework the final step of assigning subgroup labels to individuals is done by human consensus rather than by mathematical rules^13^.

The SOM was trained according to anthropometric and biochemical data; the health outcomes were excluded from the training set to prevent overfitting. A module-based approach was adopted to avoid collinearity artefacts. First, Spearman correlations were calculated for all pairs of variables. Next, the pairs of variables that were considered collinear (R^2^ > 50%) were collected into a network topology. Lastly, we used an agglomerative network algorithm to define modules of collinear variables^21^ and principal component analysis to collapse each module into a single data column.

The training set was adjusted for age and sex, centered by mean and scaled by standard deviation. The SOM was created with default settings except for smoothness = 2.0 for a more conservative fit. The quality control tests for the SOM shown in Supplementary Figure S2 (Plots A-L). We verified that every district of the map was populated (sample density ≥1,293 across the map, Plot A), the model fit was sufficient (residuals below 3 SDs, Plot B) and that the coverage of available data was high (≥92% across the map, Plot C). The map patterns were not confounded by statins (original vs. adjusted LDL, Plots D-F), by anti-hypertensives (systolic BP, Plots G-I) or by diabetic medications (glucose, Plots J-L).

Clinical diagnoses were based on three-character ICD-10 codes (International Classification of Diseases, version 10) from registers of primary care, hospital inpatients, deaths and self-reported medical conditions. Combinations of ICD-10 codes for cardiometabolic diseases are described in Supplementary Table S1. Rheumatoid arthritis, dementia and cancer were included as examples of non-cardiometabolic diseases. Cancer cases were identified using ICD-9 and ICD-10 codes from the cancer registry. The first occurrence of a disease at or before baseline was considered prevalent, new cases after baseline considered incident. Vitality status was obtained from mortality registers censored to 26^th^ April 2020.

Associations with prevalent outcomes were modelled by logistic regression and incident outcomes by Cox regression. Both model types were adjusted for age, sex and assessment center. One subgroup was chosen as the reference and the other subgroups were compared against the reference one-by-one. Cardiometabolic multimorbidity was defined as having at least two out of the four conditions (IHD, stroke, diabetes or hypertension).

Observed multimorbidity was evaluated against simulated null distributions of random co-occurrence of diseases. Firstly, a binary table was created where participants were organized as rows and diseases as columns. To obtain a random sample, the binary columns were randomly shuffled, the aggregate disease tallies were counted for each row and the proportion of rows with a disease tally greater than one was recorded. The process was repeated 10,000 times to create the null distribution. The P-value was estimated by comparing the non-shuffled proportion of multimorbidity against the null distribution. Confidence intervals were estimated similarly, except with bootstrapping instead of permutations applied to the binary table. Statistical analyses were conducted with Stata (version 16.0, College Station, TX, StataCorp LP) and R v3.5.0 (URL: https://www.R-project.org/) with the Numero library v1.4^13^.

## Results

### Correlation structure between metabolic variables

The characteristics of the study population are listed in Table 1. The mean age was 57 years (SD 8 years), most individuals were overweight (BMI mean 27.4 kg/m2, SD 4.8 kg/m2) and 20,094 (6.1%) individuals died during a mean follow-up of 10.8 years. We investigated 51 metabolic variables (34 biochemical, 15 anthropometric and two blood pressures) that were reduced to 33 SOM inputs based on collinearity (details in Methods, see also Supplementary Figure S3). The final correlation structure is shown in Figure 1.

**Table 1.** Characteristics and sex differences within the study population. Abbreviations: LDL low density lipoprotein (LDL), high density lipoprotein (HDL), systolic blood pressure (SBP), diastolic blood pressure (DBP), BMI body mass index (BMI), insulin-like growth factor-1 (IGF-1), sex hormone binding globulin (SHBG).

|  | All | Men | Women |  |
| --- | --- | --- | --- | --- |
| | Mean (SD) | Mean (SD) | Mean (SD) | $P^1_{sex}$ |
| Age (years) | 56.89 (7.99) | 57.13 (8.09) | 56.69 (7.89) | 7.5E-55 |
| BMI (kg/m<sup>2</sup>) | 27.41 (4.75) | 27.83 (4.22) | 27.04 (5.14) | <1.0E-300 |
| SBP (mmHg) | 140.2 (19.65) | 143.2 (18.47) | 137.6 (20.27) | <1.0E-300 |
| DBP (mmHg) | 82.28 (10.65) | 84.13 (10.52) | 80.67 (10.50) | <1.0E-300 |
| LDL direct (mmol/L) | 3.571 (0.87) | 3.489 (0.86) | 3.644 (0.87) | <1.0E-300 |
| HDL (mmol/L) | 1.45 (0.38) | 1.284 (0.31) | 1.599 (0.38) | <1.0E-300 |
| Triglycerides (mmol/L) | 1.755 (1.02) | 1.979 (1.14) | 1.56 (0.86) | <1.0E-300 |
| Lipoprotein A (nmol/L) | 44.17 (49.50) | 43.53 (49.42) | 44.72 (49.56) | 5.8E-10 |
| C-reactive protein (mg/L) | 2.596 (4.39) | 2.469 (4.41) | 2.707 (4.37) | 1.4E-57 |
| Glucose (mmol/L) | 5.12 (1.21) | 5.178 (1.37) | 5.066 (1.04) | 4.8E-125 |
| Glycated haemoglobin (mmol/mol) | 35.96 (6.51) | 36.3 (7.31) | 35.66 (5.71) | 2.1E-135 |
| Creatinine in plasma (μmol/L) | 72.37 (17.83) | 81.59 (17.95) | 64.32 (13.23) | <1.0E-300 |
| Cystatin C (mg/L) | 0.909 (0.17) | 0.943 (0.18) | 0.879 (0.16) | <1.0E-300 |
| Total protein (g/L) | 72.35 (4.03) | 72.46 (4.03) | 72.26 (4.03) | 1.9E-48 |
| Urate (μmol/L) | 309.5 (80.22) | 354.1 (71.35) | 270.6 (66.06) | <1.0E-300 |
| Urea (mmol/L) | 5.435 (1.39) | 5.634 (1.44) | 5.261 (1.32) | <1.0E-300 |
| Phosphate (mmol/L) | 1.16 (0.16) | 1.117 (0.16) | 1.194 (0.15) | <1.0E-300 |
| Testosterone (nmol/L) | 6.64 (6.05) | 11.969 (3.71) | 1.12 (0.62) | <1.0E-300 |
| Oestradiol (pmol/L) | 77.45 (248.64) | 20.47 (68.84) | 127 (325.88) | <1.0E-300 |
| IGF-1 (nmol/L) | 21.398 (5.66) | 21.934 (5.53) | 20.93 (5.72) | <1.0E-300 |
| SHBG (nmol/L) | 51.82 (27.65) | 39.96 (16.79) | 62.26 (30.93) | <1.0E-300 |
| Calcium (mmol/L) | 2.38 (0.09) | 2.373 (0.09) | 2.387 (0.10) | <1.0E-300 |
| Vitamin D (nmol/L) | 49.82 (20.97) | 49.85 (21.04) | 49.8 (20.91) | 0.36 |
| Alkaline phosphatase (U/L) | 83.57 (26.48) | 81.98 (24.86) | 84.95 (27.75) | 2.8E-256 |
| Rheumatoid factor (IU/ml) | 2.195 (9.14) | 2.101 (8.88) | 2.277 (9.35) | 2.3E-09 |
| Albumin in plasma (g/L) | 45.24 (2.61) | 45.53 (2.61) | 44.98 (2.59) | <1.0E-300 |
| Gamma glutamyltransferase (U/L) | 37.47 (41.89) | 45.56 (47.92) | 30.4 (34.28) | <1.0E-300 |
| Alanine aminotransferase (U/L) | 23.52 (14.01) | 27.27 (14.99) | 20.24 (12.19) | <1.0E-300 |
| Aspartate aminotransferase (U/L) | 26.19 (10.63) | 28.14 (11.40) | 24.48 (9.59) | <1.0E-300 |
| Bilirubin (μmol/L) | 9.145 (4.42) | 10.3 (4.87) | 8.135 (3.71) | <1.0E-300 |
| Albumin in urine (mg/L) | 9.231 (70.41) | 12.32 (87.42) | 6.521 (50.86) | 4.1E-116 |
| Sodium in urine (mmol/L) | 76.25 (43.57) | 87.46 (45.1) | 66.39 (39.63) | <1.0E-300 |
| Creatinine in urine (μmol/L) | 8818 (5744.03) | 10776 (6036.53) | 7100 (4863.06) | <1.0E-300 |
| Maximum follow-up (year)<sup>2</sup> | 11.01 (1.48) | 10.92 (1.63) | 11.09 (1.31) | 8.5E-203 |
|  | n (%) | n (%) | n (%) | P<sup>1</sup><sub>sex</sub> |
| IHD, prevalent | 17649 (5.35) | 12358 (8.04) | 5291 (3.00) | <1.0E-300 |
| IHD, incident<sup>3</sup> | 13690 (4.47) | 8900 (6.41) | 4790 (2.86) | <1.0E-300 |
| Stroke prevalent | 5169 (1.57) | 3101 (2.02) | 2068 (1.17) | 8.4E-73 |
| Stroke, incident<sup>3</sup> | 39137 (1.23) | 2315 (1.57) | 1602 (0.94) | 6.2E-50 |
| Diabetes, prevalent | 15558 (4.72) | 9944 (6.47) | 5614 (3.19) | 2.0E-260 |
| Diabetes, incident<sup>3</sup> | 7792 (2.53) | 4590 (3.25) | 3202 (1.92) | 2.2E-112 |
| Hypertension, prevalent | 33760 (10.23) | 18317 (11.91) | 15443 (8.77) | 7.2E-153 |
| Hypertension, incident<sup>3</sup> | 47404 (16.17) | 25600 (19.09) | 21804 (13.70) | 2.7E-300 |
| Rheumatoid arthritis, prevalent | 4338 (1.31) | 1413 (0.92) | 2925 (1.66) | 6.9E-81 |
| Rheumatoid arthritis, incident | 2040 (0.64) | 715 (0.48) | 1325 (0.78) | 3.8E-29 |
| Dementia, prevalent | 330 (0.10) | 200 (0.13) | 130 (0.07) | 1.4E-06 |
| Dementia, incident<sup>3</sup> | 2496 (0.78) | 1457 (0.97) | 1039 (0.60) | 7.1E-24 |
| Cancer, prevalent | 24438 (7.41) | 7874 (5.12) | 16564 (9.40) | <1.0E-300 |
| Cancer, incident<sup>3</sup> | 21745 (7.17) | 11459 (7.91) | 10285 (6.49) | 1.9E-39 |
| Died during follow-up<sup>2</sup> | 18481 (5.74) | 11302 (7.53) | 7179 (4.18) | <1.0E-300 |

<sup>1</sup>Linear (continuous variables) or logistic regression (binary variables) adjusted for age
<sup>2</sup>Censored on 26<sup>th</sup> April 2020
<sup>3</sup>Censored on 31<sup>st</sup> December 2016

|  | All | Men | Women |  |
| --- | --- | --- | --- | --- |
|  | Mean (SD) | Mean (SD) | Mean (SD) | P<sup>1</sup><sub>sex</sub> |
| Age (years) | 56.89 (7.99) | 57.13 (8.09) | 56.69 (7.89) | 7.5E-55 |
| BMI (kg/m<sup>2</sup>) | 27.41 (4.75) | 27.83 (4.22) | 27.04 (5.14) | <1.0E-300 |
| SBP (mmHg) | 140.2 (19.65) | 143.2 (18.47) | 137.6 (20.27) | <1.0E-300 |
| DBP (mmHg) | 82.28 (10.65) | 84.13 (10.52) | 80.67 (10.50) | <1.0E-300 |
| LDL direct (mmol/L) | 3.571 (0.87) | 3.489 (0.86) | 3.644 (0.87) | <1.0E-300 |
| HDL (mmol/L) | 1.45 (0.38) | 1.284 (0.31) | 1.599 (0.38) | <1.0E-300 |
| Triglycerides (mmol/L) | 1.755 (1.02) | 1.979 (1.14) | 1.56 (0.86) | <1.0E-300 |
| Lipoprotein A (nmol/L) | 44.17 (49.50) | 43.53 (49.42) | 44.72 (49.56) | 5.8E-10 |
| C-reactive protein (mg/L) | 2.596 (4.39) | 2.469 (4.41) | 2.707 (4.37) | 1.4E-57 |
| Glucose (mmol/L) | 5.12 (1.21) | 5.178 (1.37) | 5.066 (1.04) | 4.8E-125 |
| Glycated haemoglobin (mmol/mol) | 35.96 (6.51) | 36.3 (7.31) | 35.66 (5.71) | 2.1E-135 |
| Creatinine in plasma (μmol/L) | 72.37 (17.83) | 81.59 (17.95) | 64.32 (13.23) | <1.0E-300 |
| Cystatin C (mg/L) | 0.909 (0.17) | 0.943 (0.18) | 0.879 (0.16) | <1.0E-300 |
| Total protein (g/L) | 72.35 (4.03) | 72.46 (4.03) | 72.26 (4.03) | 1.9E-48 |
| Urate (μmol/L) | 309.5 (80.22) | 354.1 (71.35) | 270.6 (66.06) | <1.0E-300 |
| Urea (mmol/L) | 5.435 (1.39) | 5.634 (1.44) | 5.261 (1.32) | <1.0E-300 |
| Phosphate (mmol/L) | 1.16 (0.16) | 1.117 (0.16) | 1.194 (0.15) | <1.0E-300 |
| Testosterone (nmol/L) | 6.64 (6.05) | 11.969 (3.71) | 1.12 (0.62) | <1.0E-300 |
| Oestradiol (pmol/L) | 77.45 (248.64) | 20.47 (68.84) | 127 (325.88) | <1.0E-300 |
| IGF-1 (nmol/L) | 21.398 (5.66) | 21.934 (5.53) | 20.93 (5.72) | <1.0E-300 |
| SHBG (nmol/L) | 51.82 (27.65) | 39.96 (16.79) | 62.26 (30.93) | <1.0E-300 |
| Calcium (mmol/L) | 2.38 (0.09) | 2.373 (0.09) | 2.387 (0.10) | <1.0E-300 |
| Vitamin D (nmol/L) | 49.82 (20.97) | 49.85 (21.04) | 49.8 (20.91) | 0.36 |
| Alkaline phosphatase (U/L) | 83.57 (26.48) | 81.98 (24.86) | 84.95 (27.75) | 2.8E-256 |
| Rheumatoid factor (IU/ml) | 2.195 (9.14) | 2.101 (8.88) | 2.277 (9.35) | 2.3E-09 |
| Albumin in plasma (g/L) | 45.24 (2.61) | 45.53 (2.61) | 44.98 (2.59) | <1.0E-300 |
| Gamma glutamyltransferase (U/L) | 37.47 (41.89) | 45.56 (47.92) | 30.4 (34.28) | <1.0E-300 |
| Alanine aminotransferase (U/L) | 23.52 (14.01) | 27.27 (14.99) | 20.24 (12.19) | <1.0E-300 |
| Aspartate aminotransferase (U/L) | 26.19 (10.63) | 28.14 (11.40) | 24.48 (9.59) | <1.0E-300 |

<sup>1</sup>Linear (continuous variables) or logistic regression (binary variables) adjusted for age
<sup>2</sup>Censored on 26<sup>th</sup> April 2020
<sup>3</sup>Censored on 31<sup>st</sup> December 2016

**Figure 1.**
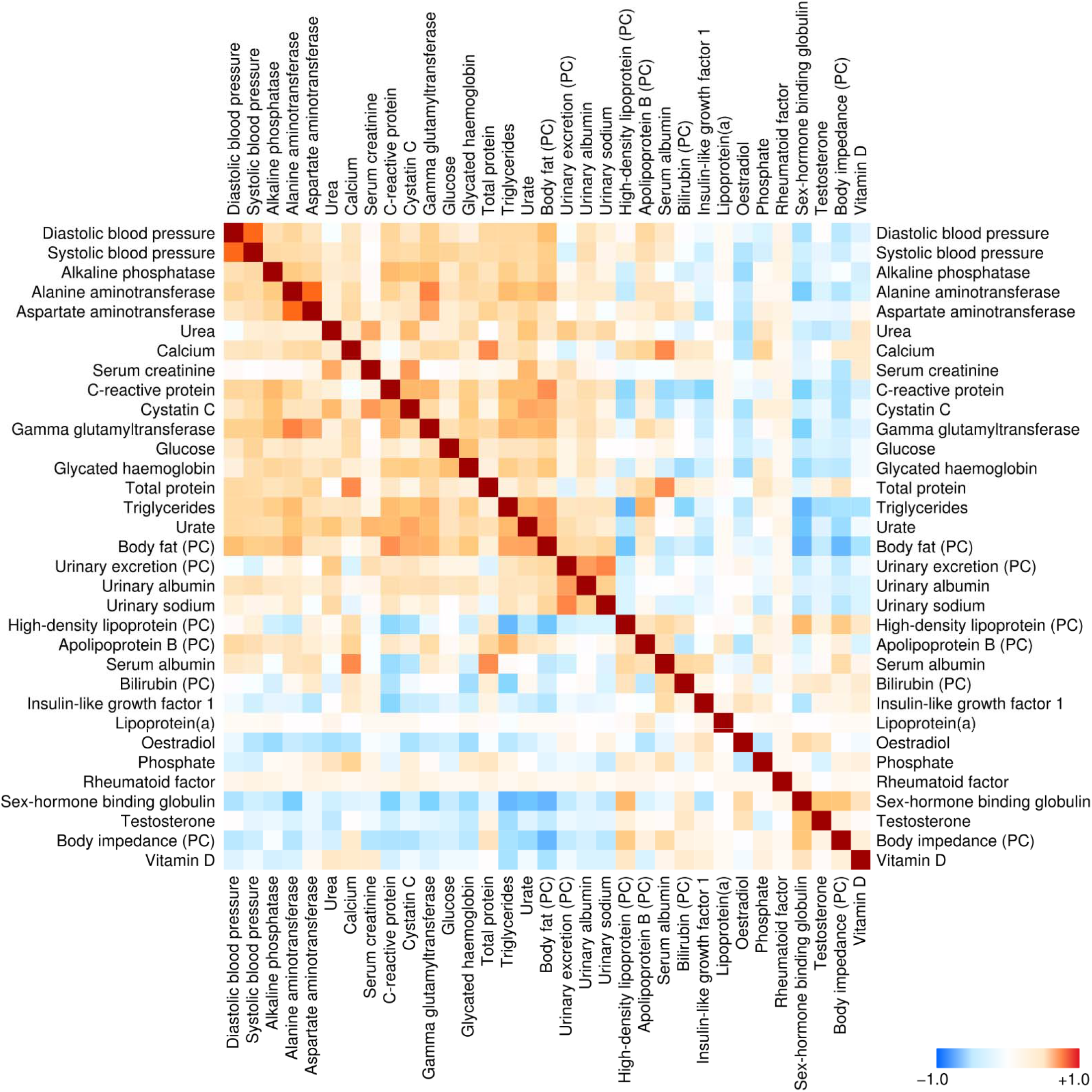
Spearman correlations between anthropometric and biochemical features that comprised the training set for the self-organizing map (adjusted for age and sex). Highly collinear variable were collapsed into the principal component score (PC) prior to correlation analysis.

### Primer on the self-organizing map

The concept of the SOM is illustrated in Figure 2. Each participant is represented by their individual preprocessed metabolic profile (Figure 2A, 33 input dimensions). The Kohonen algorithm^20^ is applied to project the high-dimensional input data onto the vertical and horizontal coordinates (two-dimensional layout in Figure 2B). On the scatter plot, proximity between two participants means that their full multivariable input data are similar as well (Figure 2C). However, scatter plots are cumbersome for large datasets and difficult to interpret in the absence of distinct clusters. The SOM circumvents these challenges by dividing the plot area into districts. To show statistical patterns, each district is colored according to the average value of a single biomarker or, in the case of morbidity, the local prevalence or incidence of a disease (Figure 2D,E). The connection between proximity on the canvas and similarity of full profile works the same way on the SOM as it does on a scatter plot. Therefore, selecting a region on the SOM is the same as selecting a subgroup of individuals with mutually similar profiles of input data (Figure 2F).

**Figure 2.**
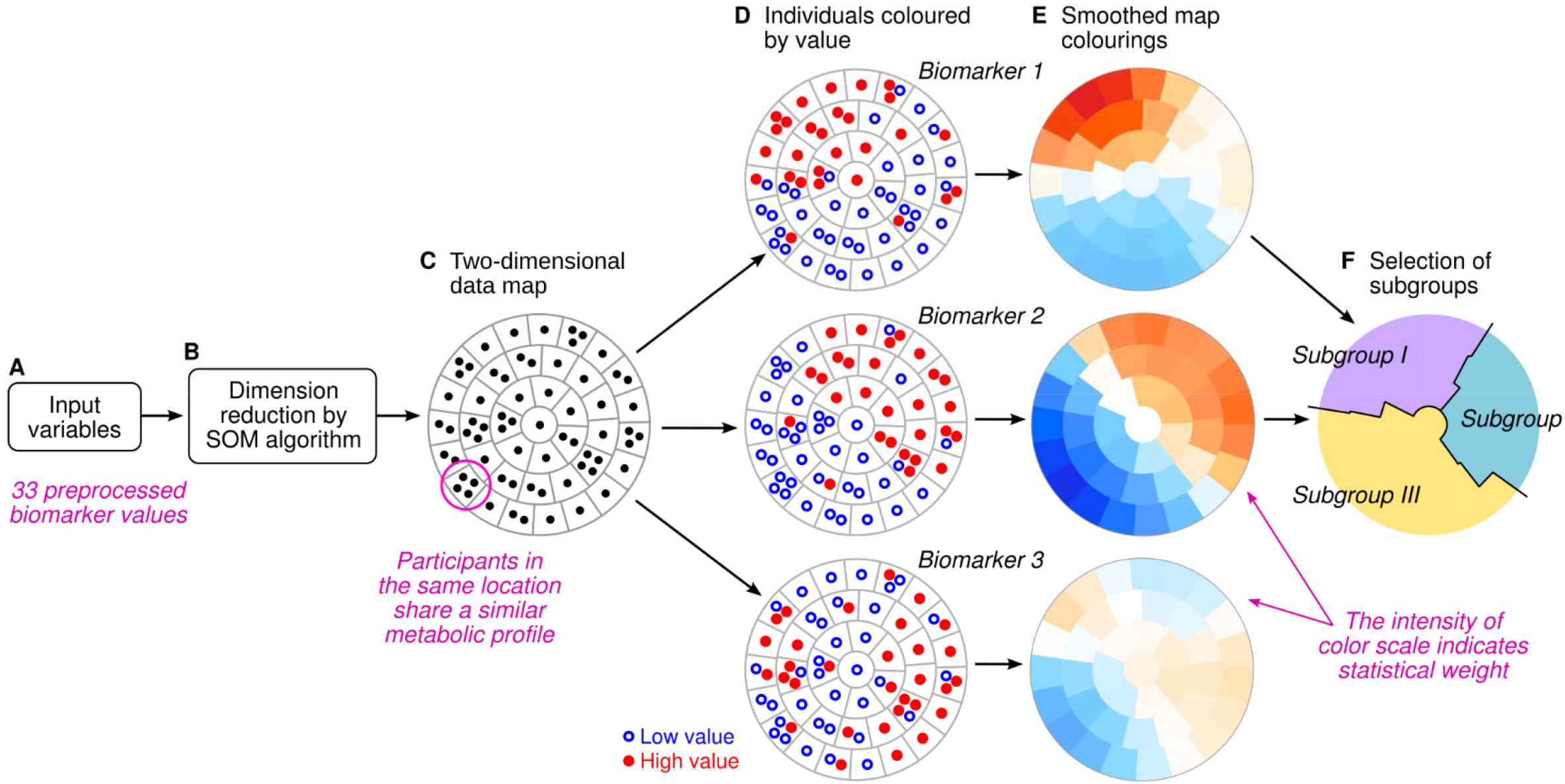
Schematic illustration of the subgrouping procedure. We used the self-organizing map (SOM) algorithm to project high-dimensional data onto a two-dimensional canvas that is divided into districts (**A-C**). The data points can be colored based on the observed values of any variable (**D**). In this study, the statistical weight of regional patterns was encoded in smoothed pseudo-colour representations of the observed values (**E**). The map colorings were used as visual guides to assign map districts and the participants therein into mutually exclusive subgroups (**F**).

### Metabolic subgroups

IHD is the most common global cause for death^22^ and causally connected to lipoproteins^23^. For this reason, we used the patterns of the apolipoprotein B module, triglycerides and the HDL module as the starting point for subgrouping (Figure 3A,G,M). We identified map regions that captured the characteristic combinations of features for individuals that had the highest apolipoprotein B score (Subgroup I, top-left part of Figure 3A-F), elevated triglycerides (Subgroups II and III, bottom-left quadrant of Figure 3G-L), and the highest HDL score (Subgroup IV, top part of Figure 3M-P).

**Figure 3.**
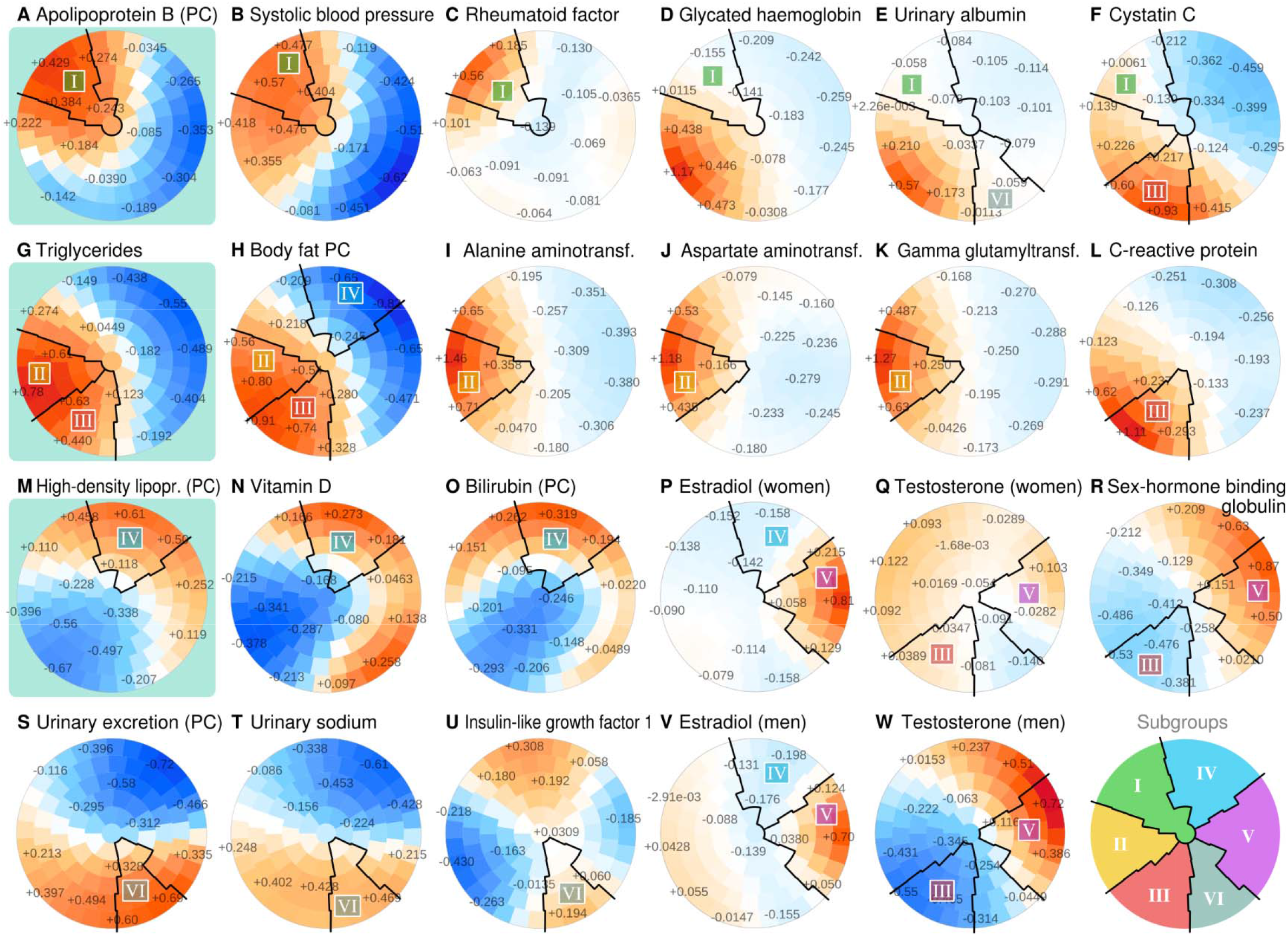
The SOM subgrouping procedure applied to the UK Biobank. In each plot, the same participants reside in the same district. The colors of the districts indicate the regional deviation from the global mean, with color intensity adjusted according to how much the variable contributed to the structure of the map. The numbers on the districts indicate the smoothed mean Z-score of th participants.

Subgroup I was characterized by the combination of high apolipoprotein B score (Figure 3A), high systolic blood pressure (Figure 3B), high rheumatoid factor (Figure 3C) and adequate glycemic control (Figure 3D). Biomarkers of kidney disease were not elevated (Figure 3E,F). The second and third subgroups featured elevated triglycerides (Figure 3G) and high body fat score (Figure 3H), however, Subgroup II was characterized by high liver enzymes (Figure 3I-K) whereas Subgroup III had higher C-reactive protein (Figure 3L). The highest HDL module scores (Subgroup IV) were observed together with the highest vitamin D (Figure 3N) and bilirubin (Figure 3O) and low estradiol (Figure 3P,V). These individuals were the leanest (Figure 3H).

The highest estradiol values were observed on the left side (Subgroup V, Figure 3P,V) and Subgroup V also showed the highest testosterone in men (Figure 3W) and sex-hormone binding globulin for both sexes (Figure 3R). Sex dimorphism was pronounced; estradiol was 5-fold higher in women, and testosterone was 10-fold higher in men and we verified that the relative SOM patterns for women under and over the age of 51^24^ were not disrupted by menopause (Supplementary Figure S4). The map area at the bottom (Subgroup VI) was characterized by high urinary excretion biomarkers without albuminuria (Figure 3E,S) and these individuals had higher insulin-like growth factor Z-scores compared to the neighboring Subgroups III and V (Figure 3U).

Succinct descriptive labels based on selected biomarkers were assigned to the subgroups for easier reading (Figure 4). Unadjusted map colorings in physical units are included in Supplementary Figures S5 and S6. Numerical descriptions of the subgroups are available in Supplementary Table S2.

**Figure 4.**
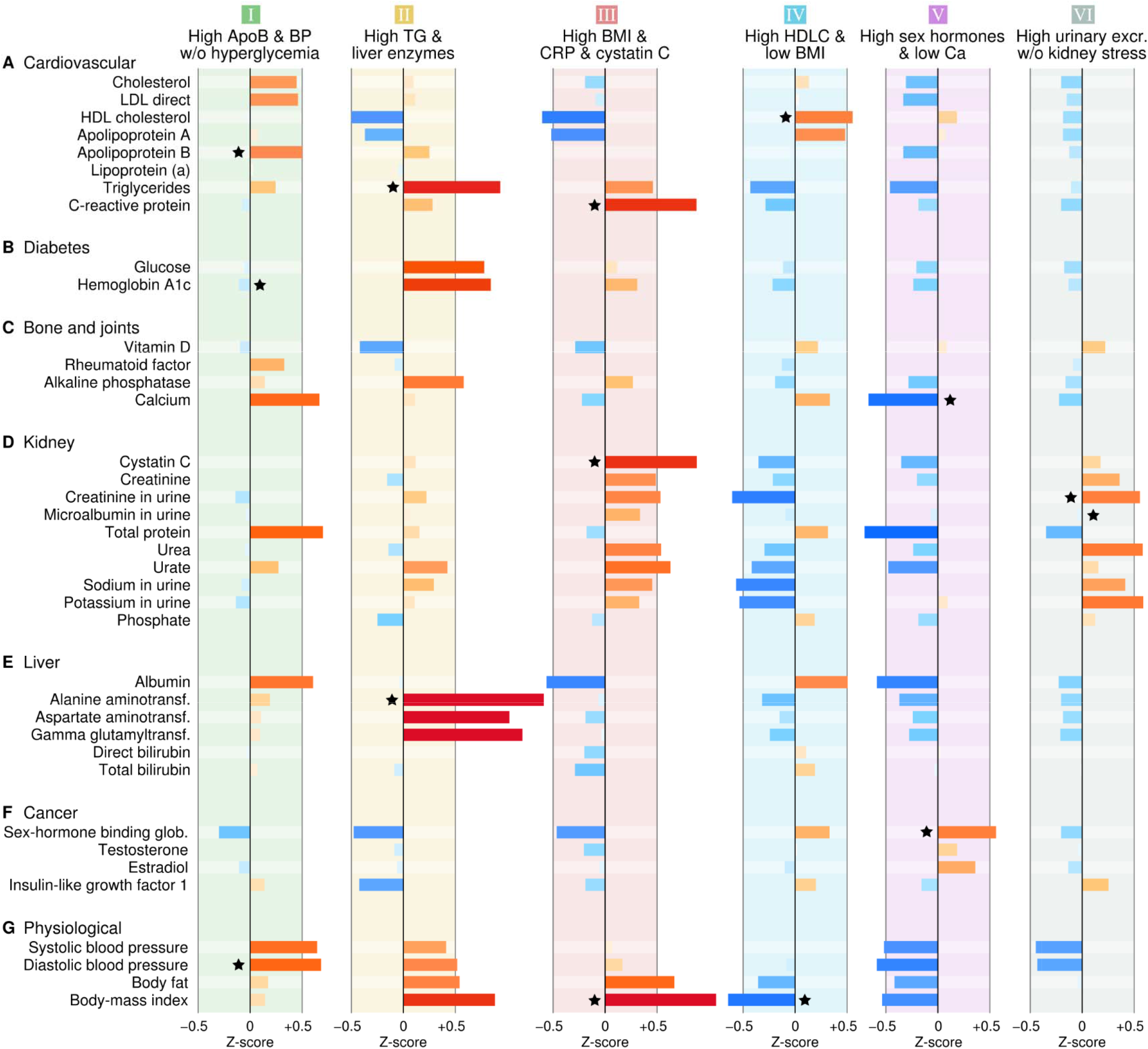
Mean metabolic profiles for SOM subgroups normalized by population SD. The bars are colored according to the direction and magnitude of the deviation from the population mean. The black stars indicate characteristic features that were selected for simplified naming of the subgroups.

### Disease prevalence and incidence by subgroup

The highest prevalence of IHD was observed in Subgroup III (Figure 5A). Diabetes prevalence varied the most across the map with small percentages for Subgroups IV and V, but substantially higher in Subgroups II and III (Figure 5B). The pattern for hypertension was close to that of diabetes (Figure 5C), but there were also individuals in Subgroup I who had hypertension (see also blood pressure in Figure 4G). The prevalence of rheumatoid arthritis, dementia and cancer was higher in Subgroup III (Figure 5D-F). Subgroup IV was associated with the lowest overall burden of disease and was chosen as the control subgroup. The subgroups were similar with respect to age, sex and follow-up time (Figure 5U-X).

**Figure 5.**
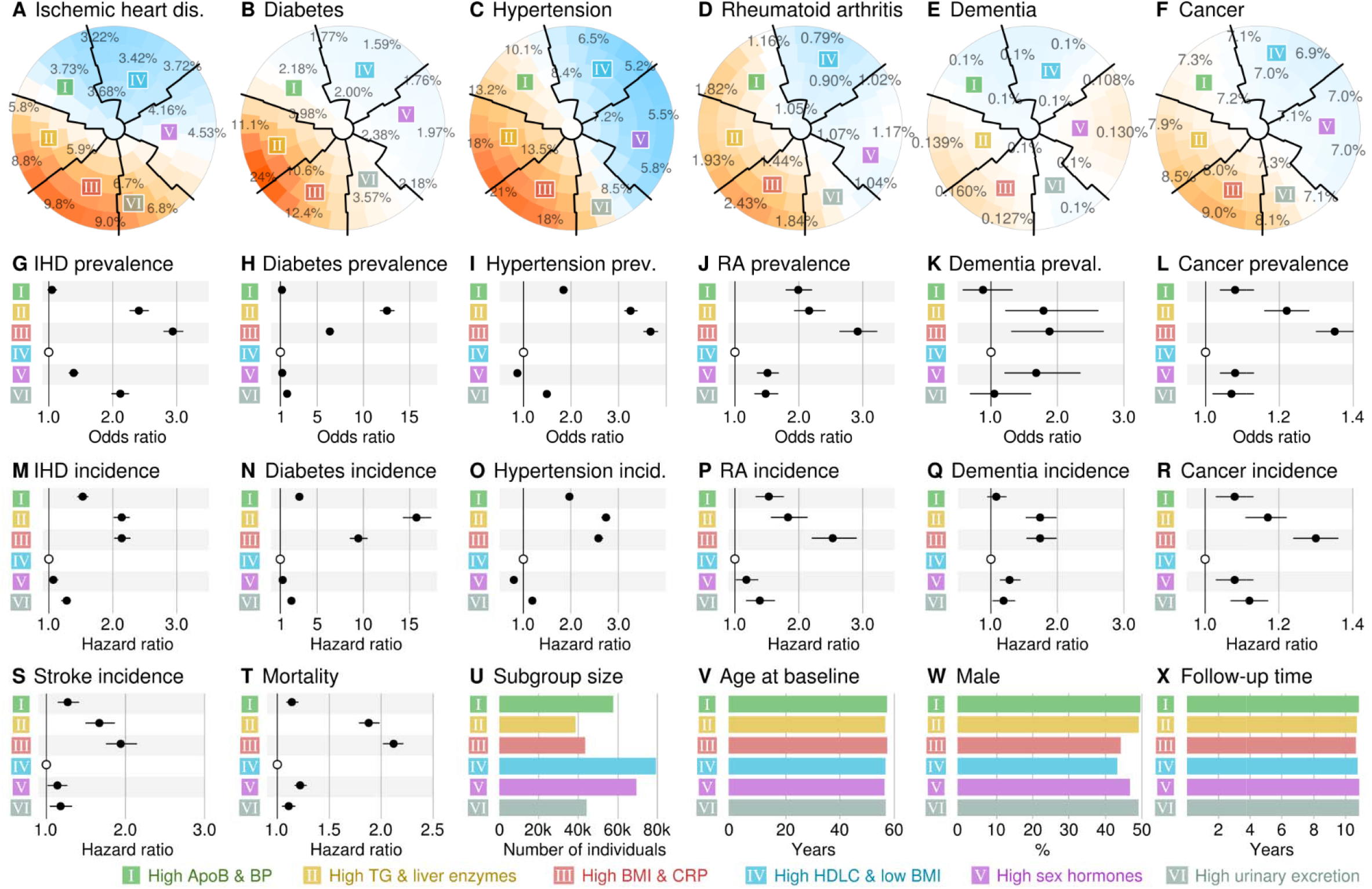
Comparison of morbidity between the SOM subgroups. Percentage of individuals with a disease at baseline across the map districts (**A-F**). Odds ratios for disease prevalence across subgrup based on logistic regression adjusted for age, sex and assessment center (**G-L**). Hazard ratios for incident disease or mortality based on Cox regression adjusted for age, sex and assessment center (**M-T**). Maximum follow-up time available across any clinical end-point (**X**).

Odds and hazard ratios of diseases between the subgroups are shown in Figure 5G-T and confidence intervals and P-values are available in Supplementary Tables S3 and S4. Subgroup III was associated with the highest prevalence of ischemic heart disease (7.5%, OR = 2.9), hypertension (19.3%, OR = 3.7), rheumatoid arthritis (2.3%, OR = 2.9) and cancer (9.1%, OR = 1.4). High incidence was observed for IHD (9.6 per 1000 person years, HR = 2.1) and the highest incidence for rheumatoid arthritis (1.6, HR = 2.53), cancer (12.8, HR = 1.3), stroke (2.6, HR = 1.9) and mortality (13.4, HR = 2.1).

The prevalence of diabetes was the highest in Subgroup II at 16.7% (OR = 12.6) and the incidence was 14.3 per 1000 person years (HR = 15.8). The incidence of ischemic heart disease in Subrgoup II was the same as in Subgroup III (9.6 vs. 9.7, P > 0.05). There were no differences in the prevalence of dementia (0.13% vs. 0.14%, P > 0.05) or the incidence of dementia (1.4 vs. 1.5, P > 0.05) between Subgroups II and III.

### Metabolic syndrome and multimorbidity

The metabolic syndrome (MetS) was developed to capture synergistic features associated with high cardiovascular risk^25,26^. The SOM patterns for MetS classification (NCEP ATP III) are shown in Figure 6A-F and numerical results are available in Supplementary Table S5. High MetS prevalence was observed in Subgroup II (64.2%) and Subgroup III (57.8%) and the lowest in Subgroup IV (5.7%).

**Figure 6.**
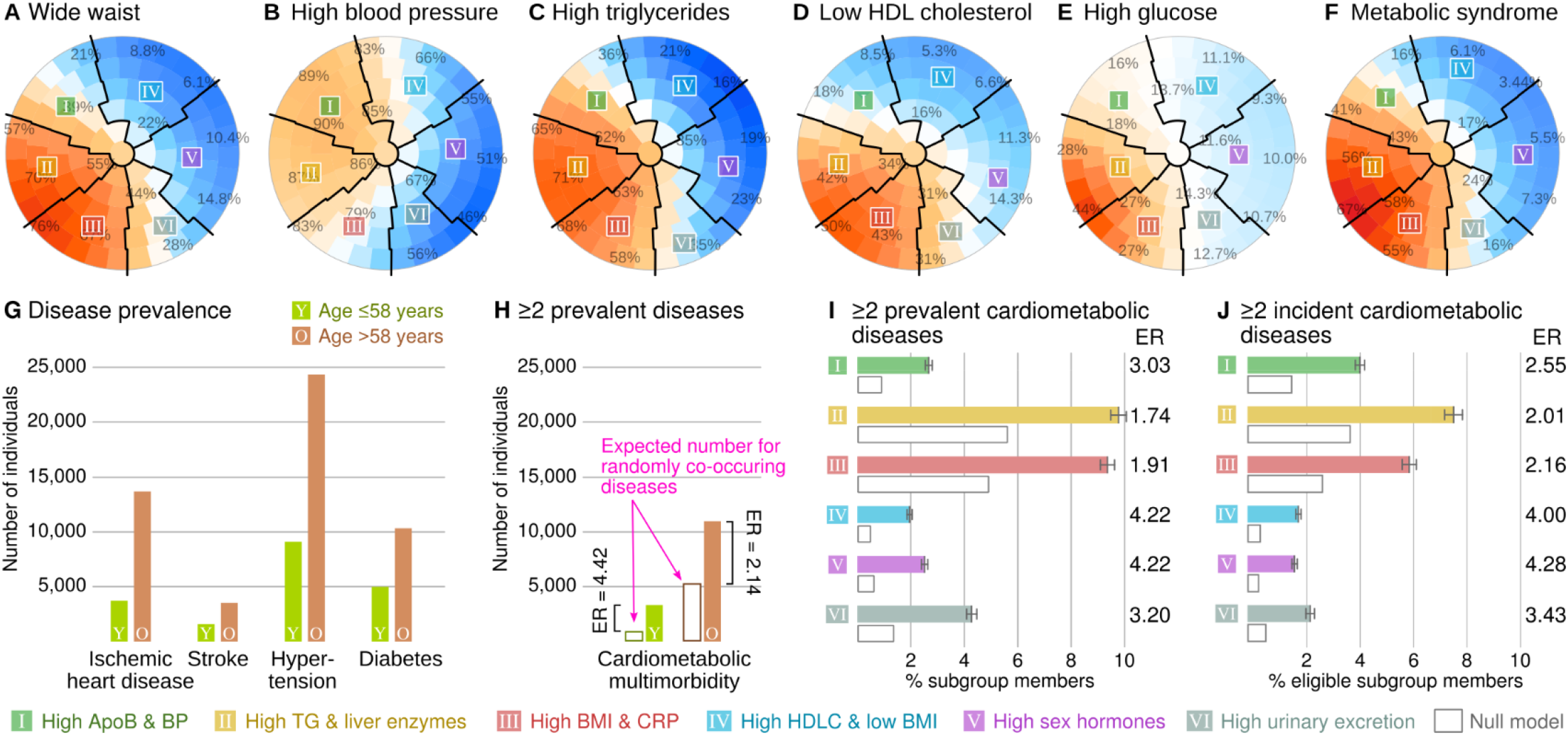
The metabolic syndrome (MetS) and multimorbidity. MetS was defined according to the NCEP ATP III criteria that include five components (**A-E**, the percentages in the plots indicate the proportion of individuals that satisfy a criterion) and subsequent binary classification for those with ≥3 points (**F**). The participants were divided into those with age ≤58 (N = 167,337 or 50.7%) and those with age >58 (N = 162,571 or 49.3%) to create two equally sized age strata (**G**). The null model represents the number of multimorbid cases if the co-occurrence of disease was random. Bars for subgroups include 95% confidence intervals (**H-J**).

The MetS combines risk factors, but we also investigated the combination of established morbidities. The burden of multimorbidity depends on the frequencies of the diseases in the population: if two diseases become more frequent, the random chance of having both increases. For example, younger individuals have fewer diseases compared to older individuals (Figure 6G, split by the median age of 58 years). This difference in disease frequencies leads to a difference in multimorbidity by mathematics alone (the null model, see Methods). However, the observed excess beyond the null model (i.e. enrichment) was greater in younger individuals (Figure 6H), which means that having one cardiometabolic disease as a young person increases the probability of having another disease more than it would for an older person.

The highest frequency of multimorbidity was observed in Subgroups II (prevalence 9.8%, incidence 7.7%) and III (prevalence 9.4%, incidence 6.1%) and the lowest in Subgroups IV (2.0%, 1.9%) and V (2.5%, 1.8%). We defined the enrichment ratio (ER) as the ratio between the observed number of individuals with ≥2 diseases versus the number predicted by the null model. Multimorbidity was enriched in all subgroups (Figure 6D,E and Supplementary Table S6), with the highest ratios observed in Subgroups IV (prevalent ER = 4.22, incident ER = 4.00), and the lowest in Subgroup II (prevalent ER = 1.74, incident ER = 2.01).

## Discussion

Metabolic dysfunction is inextricably linked with ageing demographics and the global obesity pandemic and comes with potentially grave health implications for populations and individuals alike^1–3,22^. To understand the phenomenon better, we introduced data-driven metabolic subgrouping of the UK Biobank as a model of metabolic diversity and investigated subgroup-specific prevalence and incidence of multiple clinical outcomes.

We defined six metabolic subgroups based on the SOM of the UK Biobank. The first three subgroups captured the patterns of classical IHD risk factors and the obesity pandemic (Subgroups I-III). The liver-associated Subgroup II was predictive of diabetes and IHD, which fits with the concept of fatty and insulin resistant liver as a key player in VLDL-HDL dyslipidemia, insulin resistance and type 2 diabetes^27,28^. The inflammatory and kidney stressed Subgroup III was associated with the highest mortality and overall chronic morbidity (including IHD). This pattern is also compatible with the literature^29,30^. The distinction between the liver and kidney is a notable biological insight from the SOM analysis – for example, the popular definitions of the MetS do not capture the liver-kidney spectrum^25,26^.

We identified a subgroup with elevated sex hormones (Subgroup V). These individuals had a low burden of diabetes and morbidity, which fits the Rotterdam Study^31^ and other evidence on insulin resistance^32^. Yet the Rotterdam study also reported that high estradiol in women may indicate increased diabetes risk. Furthermore, we observed multi-fold variation in absolute levels between men, women, young and old that may confound disease associations, as also noted by other studies^33–35^. Longitudinal studies with multiple time points of hormones may be necessary to understand how hormonal levels indicate and predict metabolic dysfunction.

Subgroup VI was characterized by elevated serum urea, elevated serum and urine creatinine and high urinary electrolytes. There was no clear indication of kidney stress nor high morbidity. The biochemical pattern is compatible with the expected effects of habitual high-protein diet^36^. Subgroup VI may also capture a haemodynamic or a fluid balance aspect of metabolic health^37^. Incidental circumstances during sample collection is another possibility: as there is only one biochemical time point, acute illness or other stressors before the baseline visit may have confounded systemic metabolism and resulted in atypical findings for multiple affected and correlated biomarkers.

Obesity and unfavourable lifestyle are risk factors for multimorbidity^1,38,39^. However, the previous studies did not consider the confounding increase in co-occurrence when the frequency of diseases increases. We observed a synergistic enrichment for cardiometabolic multimorbidity in all subgroups. The most likely explanation is intertwined etiology, partly due to pleiotropic genetic variants and environmental exposures^40,41^ and partly due to secondary effects between the diseases themselves such as the mechanical stress on the vasculature from hypertension^42^ or toxicity from excessive glycation in diabetes^43^. Another explanation could be diagnostic procedures: if one disease is detected, it is easier to look for and establish the presence of another.

Multimorbidity enrichment was pronounced in the metabolically favorable Subgroups IV and V despite them having lower disease burden overall. The paradoxical finding means that the relative risk of co-occurring cardiometabolic disease was higher in the absence of obvious metabolic abnormality. The pattern may reflect genetic and environmental susceptibility that is independent of the typical cardiovascular risk factors but nevertheless pleiotropic to cardiometabolic diseases^44,45^. The same pattern may also arise from survival bias as people who are simultaneously affected by metabolic dysfunction and multiple morbidities tend to perish younger^46^.

The juxtaposition between the population and an individual is relevant for the aspirational goals of precision medicine^47^. The SOM provides the opportunity to connect the two domains in a meaningful way. A subgroup profile can be presented as a list of measurement values in their physical units and it is easy for human observers to verify which profile matches their own. Therefore, the SOM model is directly applicable to real-world people and the results can be communicated without mathematical abstractions. Yet a subgroup contains multiple individuals, which enables the calculation of prevalence and incidence rates as subpopulation risk estimates. Indeed, propensity scoring is already used in this manner to identify pools of representative cases within health informatics systems^48,49^. However, these methods are often presented as black boxes and thus lack the biological context that the SOM colorings can provide.

Due to the large sample size, the statistical robustness is high in this study but we urge caution when generalizing the findings of this study to other cohorts, to other ethnicities or to populations of different circumstances. The UK Biobank recruited volunteers only, thus people with less opportunity to participate due to low socio-economic status or poor health may be under-represented, however, the disease associations are compatible with other cohorts^50^. Ageing affects metabolism, but the SOM was constructed from cross-sectional data and adjusted for age, thus we are unable to provide information on longitudinal metabolic trajectories and the metabolic subgroups should not be interpreted as part of a temporal sequence.

In conclusion, the SOM subgroup modeling of the UK Biobank supported our hypothesis that metabolic diversity predicts disease diversity later in life. We also observed unexpected patterns of multimorbidity that are relevant for the identification of metabolically “healthy” individuals who might still be at elevated risk of disease. These results demonstrate how the health of an entire population can be summarized and simplified using an artificial neural network and how the subgrouping concept creates new opportunities to monitor and intervene in public health settings as more and more high-dimensional medical data become available.

## Data Availability

Data will be made available through application to the UK Biobank.

## Acknowledgments

We thank the UK Biobank participants and administrators for making this study possible.

## Funding

This work was supported by the NHMRC grant GNT1157281. MAK was supported by a research grant from the Sigrid Juselius Foundation, Finland.

## Author contributions

VPM and EH conceived the study, AM and VPM analyzed the data, all authors interpreted the results and participated in the writing of the manuscript.

## Competing interests

No conflicts of interest.

## Data and materials availability

The UK Biobank data are publicly available (https://www.ukbiobank.ac.uk/). This study was designed and implemented according to project plan #29890.

**Supplementary Figure S1**

Participant selection.

**Supplementary Figure S2**

SOM quality control. Sample density indicates the number of UK Biobank participants located within a map district (**A**). Model residuals indicate how well the SOM captures the shape of the metabolic profiles for individuals located within a map district. Values between −3 and +3 are considered acceptable quality (**B**). Data availability indicates the proportion of usable measurement values (**C**). Selected quantitative traits were adjusted for the appropriate drug effects to check if the SOM patterns were confounded by medication (**D-L**).

**Supplementary Figure S3**

Correlation modules of biomarkers. The modules were derived using an agglomerative from the pair-wise Spearman correlation network between 51 metabolic traits. First, edges with R^2^ < 50% were excluded, then an agglomerative spanning tree algorithm was applied to determine highly connected modules.

**Supplementary Figure S4**

SOM colorings for hormones, stratified by sex and the mean age of menopause. Z-scores indicate values of standardized input features as used in the SOM training (three columns of plots on the left). The measured values were not adjusted and are reported in their original measurement units. Furthermore, the map colors are calibrated in such a way that the same numerical value corresponds to the same color in each plot of a specific variable (three columns of plots on the right).

**Supplementary Figure S5**

SOM colorings for men.

**Supplementary Figure S6**

SOM colorings for women.

**Supplementary Table S1**

Diagnostic codes.

**Supplementary Table S2**

Subgroup profiles.

**Supplementary Table S3**

Subgroup disease prevalence.

**Supplementary Table S4**

Subgroup disease incidence.

**Supplementary Table S5**

Metabolic syndrome.

**Supplementary Table S6**

Multimorbidity.

## Notes

### Competing Interest Statement

The authors have declared no competing interest.

### Clinical Trial

This study is based on data from UK Biobank cohort.

### Author Declarations

UK Biobank obtained informed consent from each participant, and ethical approval was granted by the National Information Governance Board for Health and Social Care and North West Multi-centre Research Ethics Committee (11/NW/0382). The current study is approved by the UK Biobank under application number 29890.

