## Supplementary figures for "Metabolic subgroups and cardiometabolic multimorbidity in the UK Biobank"

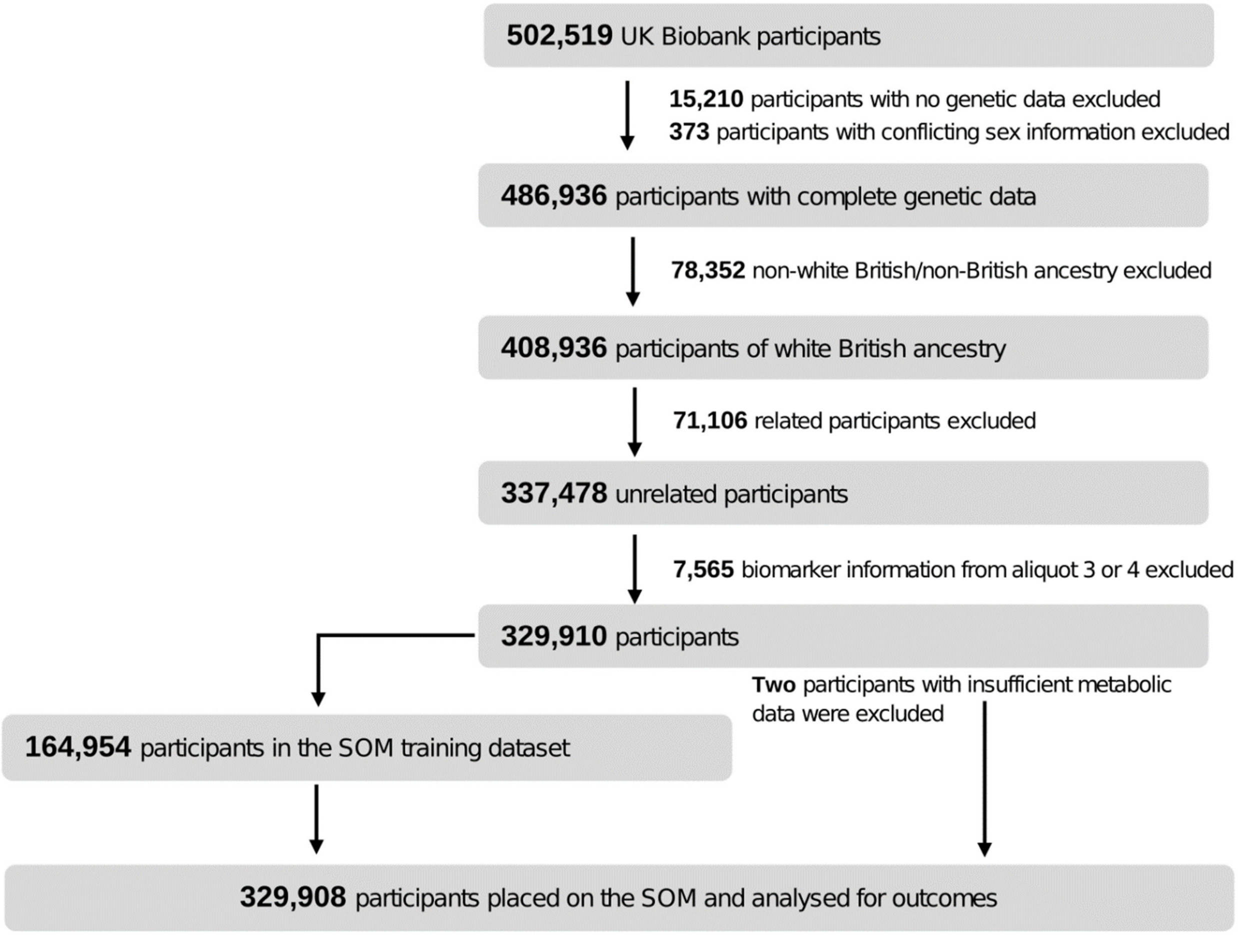


**Supplementary Figure S1**

Participant selection.

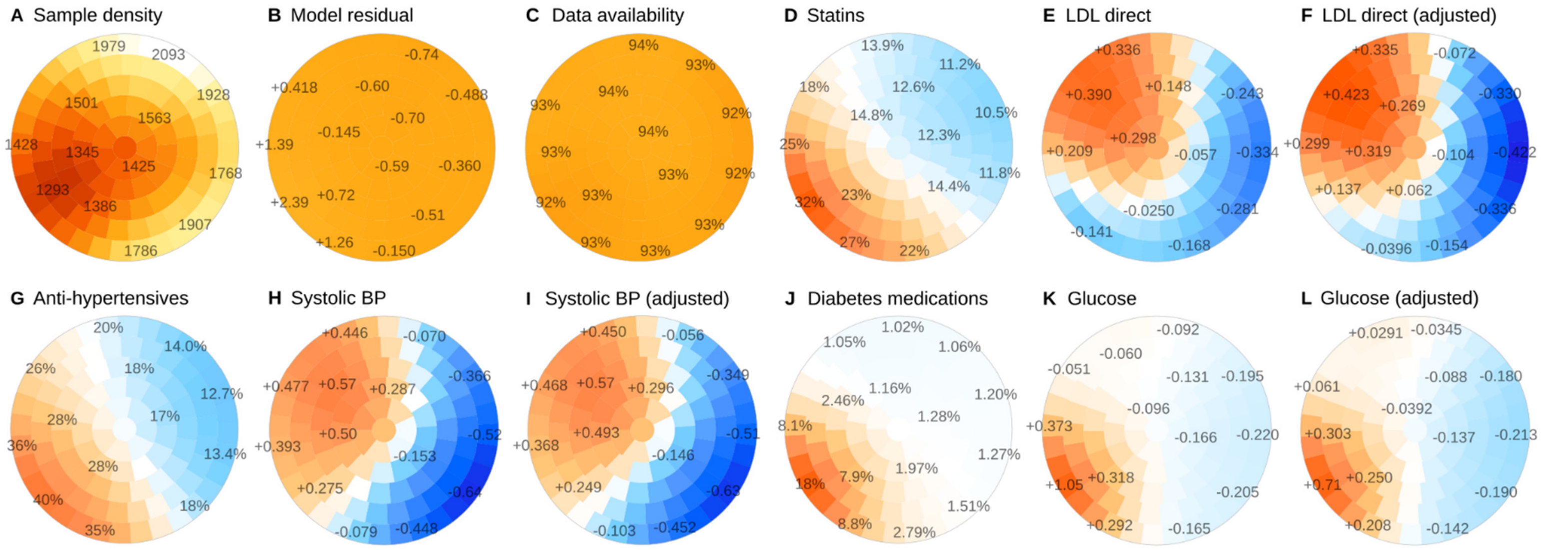


**Supplementary Figure S2**

SOM quality control. Sample density indicates the number of UK Biobank participants located within a map district (**A**). Model residuals indicate how well the SOM captures the shape of the metabolic profiles for individuals located within a map district. Values between −3 and +3 are considered acceptable quality (**B**). Data availability indicates the proportion of usable measurement values (**C**). Selected quantitative traits were adjusted for the appropriate drug effects to check if the SOM patterns were confounded by medication (**D-L**).

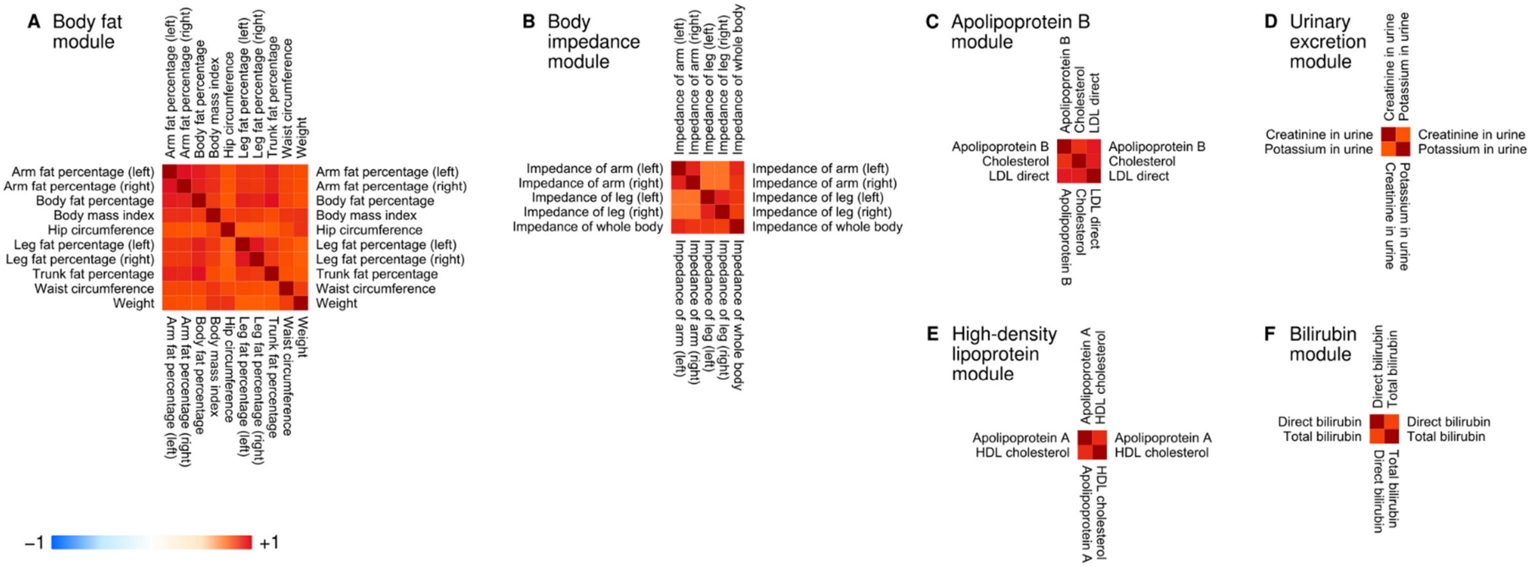


**Supplementary Figure S3**

Correlation modules of biomarkers. The modules were derived using an agglomerative from the pair-wise Spearman correlation network between 51 metabolic traits. First, edges with $R^2 < 50\%$ were excluded, then an agglomerative spanning tree algorithm was applied to determine highly connected modules.

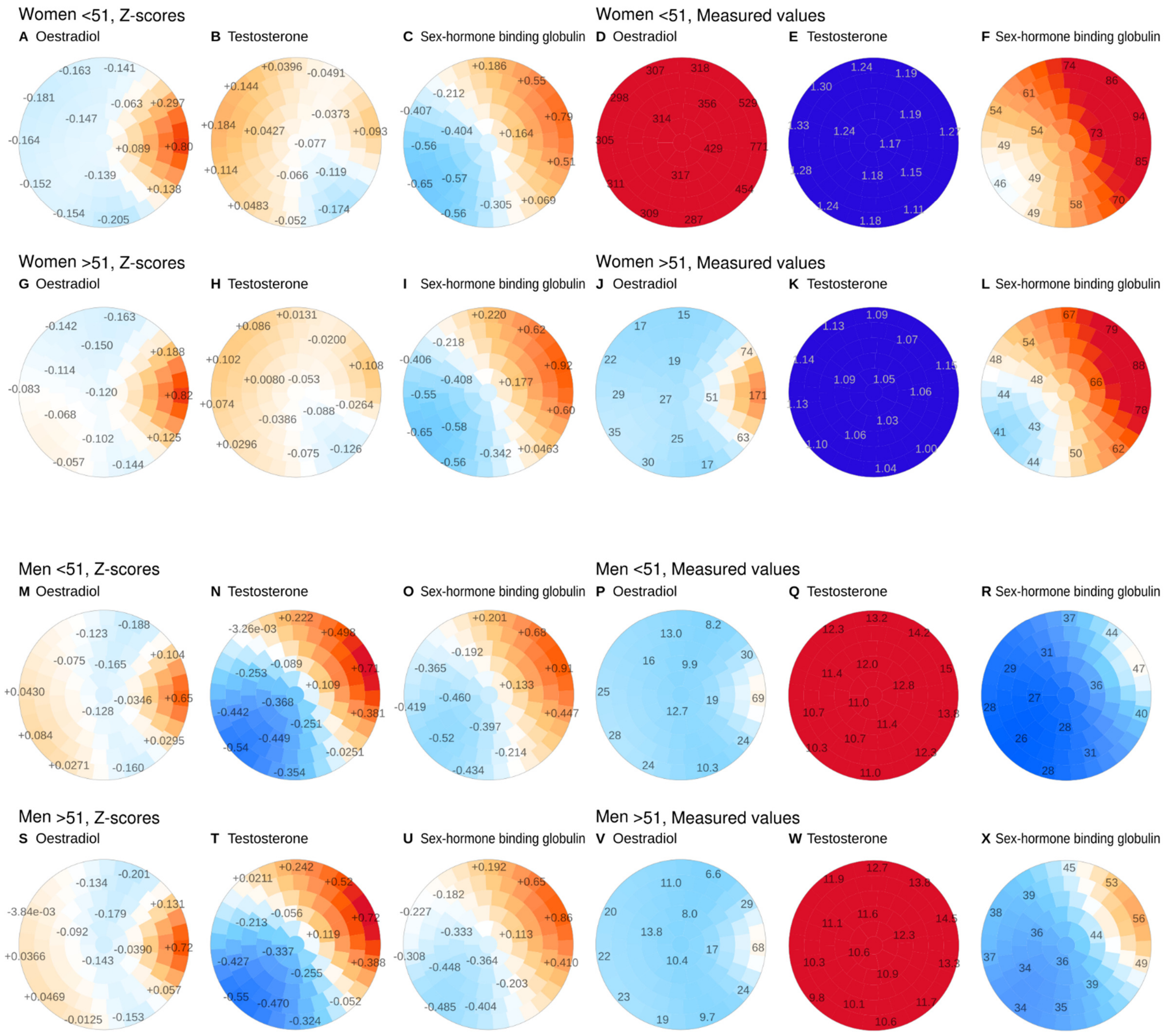


**Supplementary Figure S4**

SOM colorings for hormones, stratified by sex and the mean age of menopause. Z-scores indicate values of standardized input features as used in the SOM training (three columns of plots on the left). The measured values were not adjusted and are reported in their original measurement units. Furthermore, the map colors are calibrated in such a way that the same numerical value corresponds to the same color in each plot of a specific variable (three columns of plots on the right).
